# Altered gene expression and PTSD symptom dimensions in World Trade Center responders

**DOI:** 10.1101/2021.03.05.21252989

**Authors:** Shelby Marchese, Leo Cancelmo, Olivia Diab, Leah Cahn, Cindy Aaronson, Nikolaos P. Daskalakis, Jamie Schaffer, Sarah R Horn, Jessica S. Johnson, Clyde Schechter, Frank Desarnaud, Linda M Bierer, Iouri Makotkine, Janine D Flory, Michael Crane, Jacqueline M. Moline, Iris G. Udasin, Denise J. Harrison, Panos Roussos, Dennis S. Charney, Karestan C Koenen, Steven M. Southwick, Rachel Yehuda, Robert H. Pietrzak, Laura M. Huckins, Adriana Feder

## Abstract

Despite experiencing a significant trauma, only a subset of World Trade Center (WTC) rescue and recovery workers developed posttraumatic stress disorder (PTSD). Identification of biomarkers is critical to the development of targeted interventions for treating disaster responders and potentially preventing the development of PTSD in this population. Analysis of gene expression from these individuals can help in identifying biomarkers of PTSD.

We established a well-phenotyped sample of 371 WTC responders, recruited from a longitudinal WTC responder cohort, by obtaining blood, self-reported and clinical interview data. Using bulk RNA-sequencing from whole blood, we examined the association between gene expression and WTC-related PTSD symptom severity on (i) highest lifetime Clinician-Administered PTSD Scale (CAPS) score, (ii) past-month CAPS score, and (iii) PTSD symptom dimensions using a 5-factor model of re-experiencing, avoidance, emotional numbing, dysphoric arousal and anxious arousal symptoms. We corrected for sex, age, genotype-derived principal components and surrogate variables. Finally, we performed a meta-analysis with existing PTSD studies (total N=1,016), using case/control status as the predictor and correcting for these variables.

We identified 66 genes significantly associated with highest lifetime CAPS score (FDR-corrected p<0.05), and 31 genes associated with past-month CAPS. Our more granular analyses of PTSD symptom dimensions identified additional genes that did not reach statistical significance in our overall analysis. In particular, we identified 82 genes significantly associated with lifetime anxious arousal symptoms. Several genes significantly associated with multiple PTSD symptom dimensions and lifetime CAPS score (*SERPINA1, RPS6KA1,* and *STAT3*) have been previously associated with PTSD. Geneset enrichment of these findings has identified pathways significant in metabolism, immune signaling, other psychiatric disorders, neurological signaling, and cellular structure. Our meta-analysis revealed 10 genes that reached genome-wide significance, all of which were down-regulated in cases compared to controls (*CIRBP, TMSB10, FCGRT, CLIC1, RPS6KB2, HNRNPUL1, ALDOA, NACA, ZNF429 and COPE*). Additionally, cellular deconvolution highlighted an enrichment in CD4 T cells and eosinophils in responders with PTSD compared to controls.

The distinction in significant genes between lifetime CAPS score and the anxious arousal symptom dimension of PTSD highlights a potential biological difference in the mechanism underlying the heterogeneity of the PTSD phenotype. Future studies should be clear about methods used to analyze PTSD status, as phenotypes based on PTSD symptom dimensions may yield different gene sets than combined CAPS score analysis. Potential biomarkers implicated from our meta-analysis may help improve therapeutic target development for PTSD.

## Introduction

Posttraumatic stress disorder (PTSD) is a complex psychiatric disorder that can develop after experiencing a traumatic event. The attacks on the World Trade Center (WTC) on September 11, 2001 and their aftermath had a substantial impact on the physical and mental health of WTC rescue, recovery and clean-up workers, but only a subset developed PTSD. These differing clinical outcomes after experiencing trauma imply a role for biological and genetic influence in PTSD. Our cohort provides an unprecedented opportunity into PTSD insights, because they have been deeply phenotyped for a shared, specific trauma.

Understanding the biological mechanisms underlying PTSD will require careful dissection and analysis of many constituent symptoms and risk factors. PTSD is uniquely heterogeneous among psychiatric disorders, with complex and detailed diagnostic criteria that allow for 636,120 different combinations of symptoms^1^, and 79,794 different symptom combinations. Additional heterogeneity in PTSD stems from the type and extent of trauma. It has been well established that PTSD is a heterogenous disorder and that trauma type plays a role in differential outcomes. In the field of WTC exposures, some work has already been done to elucidate gene expression and clinical outcomes.^2–10^ Further, work by our group and others has demonstrated differential genetic heritability of PTSD according to trauma-type^11, 12^.

Identification of biomarkers will be critical to the development of targeted interventions for treating disaster responders and potentially preventing the development of PTSD in this population. Gene expression analysis from WTC responders is uniquely useful to deduce the biological heterogeneity in PTSD, given their exposure to a similar and well-documented trauma. Data on WTC-related traumatic exposures of responders analyzed here, in combination with their heterogenic clinical outcomes, makes this a critical study to understand PTSD development and chronicity after shared traumatic events. Although candidate gene expression and methylation studies have explored genes involved in canonical stress signaling pathways in PTSD, regulated by the hypothalamus-pituitary-adrenal (HPA) axis, and immune and sympathetic nervous systems, few have been able to control for length of time since exposure, nor so specifically delineate trauma type and secondary exposures such as dust cloud severity. While the WTC-related exposures experienced by rescue, recovery and clean-up workers in this cohort ranged in severity, the traumatic event –encompassing the 9/11 attacks and their aftermath– happened in a discrete, specific time window. Further, this sample is highly phenotyped with in-person clinical psychiatric evaluations, also including medical examination and laboratory testing.

The existence of this cohort and the generous participation of many responders to the WTC disaster enabled us to generate a large gene expression data set of 355 donors, to our knowledge the largest single traumatic event expression data set to date. We used the Clinician-Administered PTSD Scale (CAPS)^13^ score as a quantitative measure of PTSD symptom severity rather than a case/control definition, thus substantially increasing statistical power in this study.^14–16^ To our knowledge, ours is the first gene expression study to incorporate total CAPS scores and PTSD symptom dimensions as outcomes.

## Methods

### Participants

The WTC Health Program (WTC-HP) is a regional consortium of five clinical centers established in the greater New York City area by the Centers of Disease Control and Prevention in 2002, with the goal of providing health monitoring and treatment to WTC responders, comprising the WTC-HP General Responder Cohort^17^. We recruited participants from the WTC-HP Responder Cohort who had completed at least three periodic health monitoring visits at one of the four WTC-HP clinical centers participating in this study – Mount Sinai Medical Center, New York University, Northwell Health, and Rutgers/The State University of New Jersey – and who had provided signed consent to be contacted for research studies. Stratified random sampling was employed to ensure selection of WTC responders spanning the full range of WTC-related PTSD symptom severity, from no/minimal symptoms to severe/chronic PTSD symptom levels on the PTSD Checklist – Specific Version (PCL-S)^18^ completed during periodic health monitoring visits to the WTC-HP. Individuals with a lifetime history of chronic psychotic disorder or bipolar disorder type I, substance abuse/dependence or alcohol dependence over the prior three months, current pregnancy, acute medical illness or exacerbation of chronic medical illness, history of significant head injury or cerebrovascular accident, changes in medications or medication dosages over the prior month, or who were taking oral or regularly injected steroid medications were excluded from the study.

The study, conducted between April 2013 and September 2017, was approved by the Institutional Review Board of the Icahn School of Medicine at Mount Sinai, and all participants provided written informed consent. A total of 471 WTC responders completed in-person clinical assessments, yielding a final sample of 371 participants who met study eligibility criteria and completed study procedures, and 355 of those who had viable RNA-sequencing data (Figure 1). The mean age of participants was 54.1 (SD=8.3) years, 82% were male; ethnicity proportions are given in Table 1. The sample was composed of 40.8% police responders and 59.2% non-traditional responders (e.g., construction workers).

**Figure 1.**
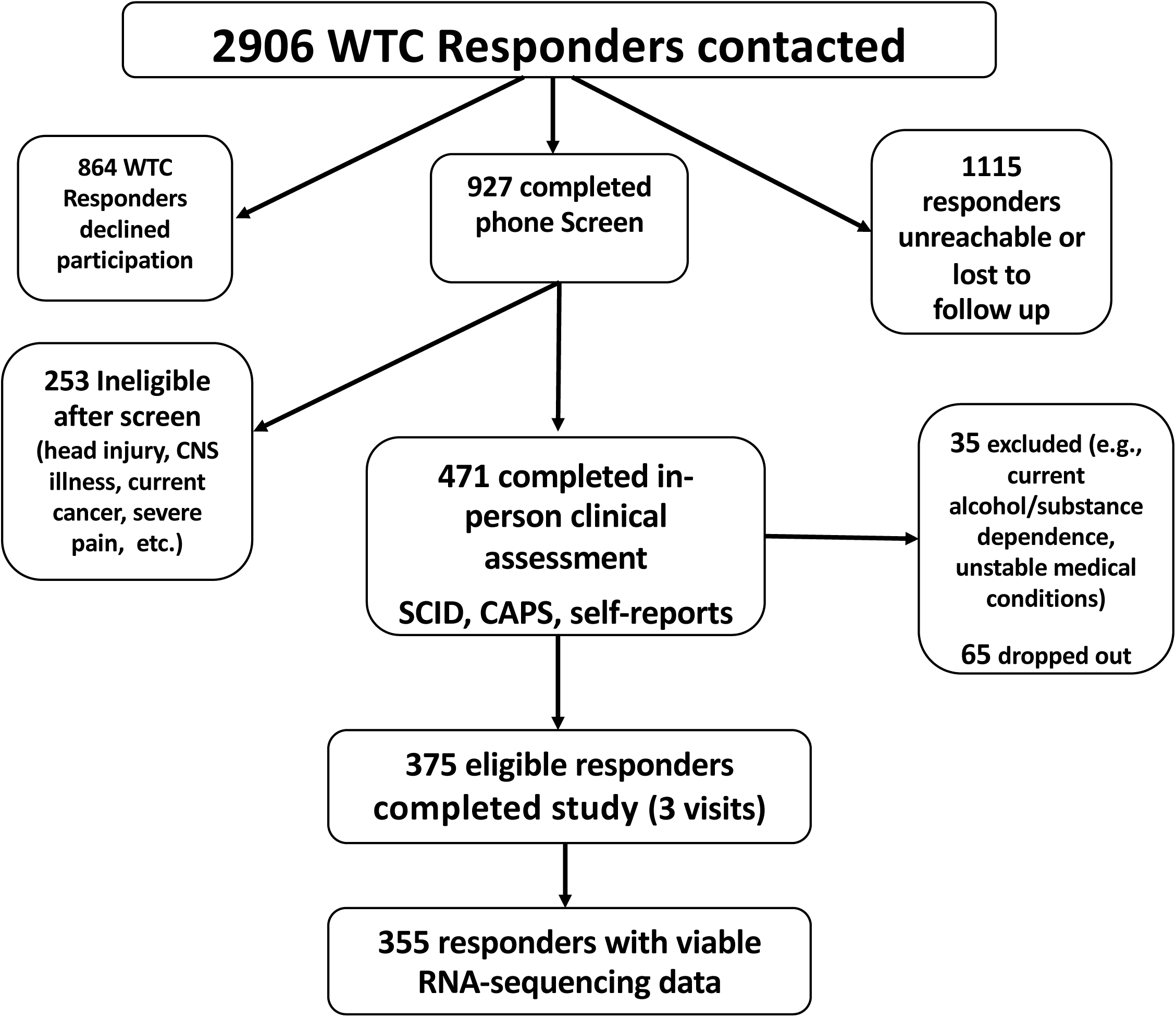
Number of World Trade Center first respondents who completed the study with viable RNA-sequencing data, N=355.

**Table 1.** Demographic information of N=355 World Trade Center first responders.

|  | Mean (SD) or N (%) |  |
| --- | --- | --- |
| Age (years) | 54.1 (8.3) | *Mean (SD)* |
| Number of years post-9/11 sample collected | 13 (2.3) | *Mean (SD)* |
| BMI | 30.1 (5.9) | *Mean (SD)* |
| Male sex | 291 (82) | *N (%)* |
| Current smoker | 22 (6.2) | *N (%)* |
| High school graduate | 301 (84.9) | *N (%)* |
| Annual income > \$90,000 | 178 (50.1) | *N (%)* |
| Responder type |  |  |
| Police responder | 145 (40.8) | *N (%)* |
| Non-traditional responder | 210 (59.2) | *N (%)* |
| Race-ethnicity |  |  |
| African American | 64 (18) | *N (%)* |
| Asian | 5 (1.4) | *N (%)* |
| Hispanic | 72 (20.3) | *N (%)* |
| Native American | 9 (2.5) | *N (%)* |
| European American (White) | 216 (60.8) | *N (%)* |
| Other | 8 (2.3) | *N (%)* |

Note: SD = standard deviation; BMI= body mass index; n=355

### Assessments

Data on 10 WTC-related exposures ^19^(e.g., exposed to human remains, received treatment for an illness/injury during WTC recovery work) was obtained from interviews and self-report questionnaires completed by participants during their first health-monitoring visit to the WTC-HP, an average of 4.3 (SD=2.7) years following 9/11/2001. In-person clinical assessments were conducted an average of 13 (SD=2.3) years following 9/11/2011. Trained Masters- or PhD-level clinical interviewers administered the Structured Clinical Interview for DSM-IV (SCID)^1^ to assess current and lifetime Axis-I psychiatric diagnoses, and CAPS^13^, lifetime and past-month versions, to assess lifetime and past-month WTC-related PTSD diagnostic status and WTC-related PTSD symptom severity. Lifetime and past-month PTSD diagnosis was defined as meeting DSM-IV criteria for WTC-related PTSD and a total score ≥ 40 on the lifetime and past-month CAPS, respectively.

On the same day as the clinical assessment, participants also completed the Childhood Trauma Questionnaire (CTQ)^20^, assessing physical, sexual, and emotional abuse, and physical and emotional neglect experienced in childhood; the Traumatic Life Events Questionnaire (TLEQ)^21^, assessing lifetime exposure to a range of traumatic events (e.g, crime, natural disaster, assault); a checklist of 15 stressful life events they might have experienced since 9/11/2011 (e.g., “lost a job/laid off/lost income”, “divorced from spouse”, “had debt trouble”), modified from the Diagnostic Interview Schedule (DIS) Disaster Supplement^22^; and a health questionnaire asking which medical conditions they had ever been diagnosed with^23^, modified to add common WTC-related conditions (asthma or chronic respiratory condition, chronic rhinitis or sinusitis, sleep apnea, or acid reflux). Participants additionally completed a history and physical examination conducted by a licensed nurse practitioner, as well as routine laboratory testing, to rule out acute medical illness or exacerbation of chronic medical illness.

Among the 355 participants, 108 were determined to have met DSM-IV criteria for lifetime WTC-related PTSD, with 53 of them still meeting past-month PTSD criteria. The heterogeneity of PTSD confers some problems when attempting to analyze the disorder by case/control status alone. Case/control analyses do not fully capture the symptom complexity of the disorder, resulting in poor genomic modeling. Similarly, while overall PTSD symptom severity is a better quantitative measurement, it does not fully capture variability across PTSD symptomatology on a useful clinical level.^24^ To address this variability, we examined five symptom dimensions (re-experiencing, avoidance, emotional numbing, dysphoric arousal and anxious arousal symptoms), assessed with the CAPS to more accurately examine the heterogeneity of PTSD symptomatology (Figure 2).^25^

**Figure 2.**
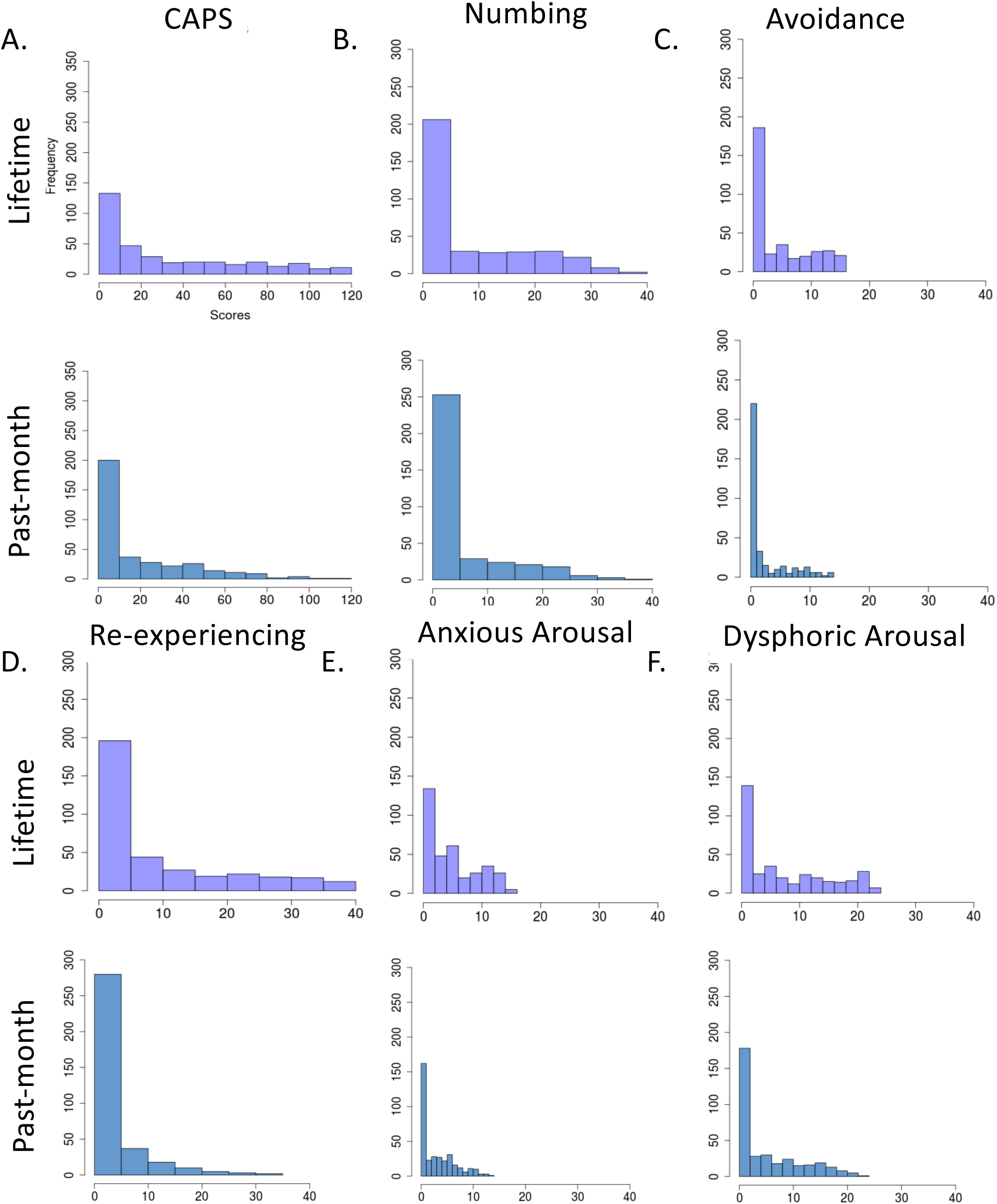
Distribution of lifetime and past-month severity scores for (A) CAPS, (B) numbing, (C) avoidance, (D) re-experiencing, (E) anxious arousal and (F) dysphoric arousal. PTSD symptom dimensions are scored from 0-50, and CAPS is an aggregate of that score. Total N=355 World Trade Center responders.

### Blood sample collection and RNA extraction

Participants were instructed to fast after midnight and underwent collection of blood samples between 8:00 and 10:00 am. Total RNA was purified from whole blood collected in PAXgene blood RNA tubes (Qiagen, Germantown, MD, USA) using a PAXgene blood RNA kit IVD (Qiagen, Germantown, MD, USA). Total RNA concentration and quality were estimated using a NanoDrop 200c spectrophotometer according to the manufacturer instructions (Thermo Fisher Scientific, Waltham, MA, USA). Samples with an optical density ratio 260/280 superior or equal to 1.8 passed the quality control. Total RNA concentration and quality were also estimated using an Agilent RNA 6000 nano kit and an Agilent 2100 bioanalyzer according to the manufacturer instructions (Agilent Technologies, Santa Clara, CA, USA). Samples with an RNA Integrity Number superior or equal to 7 passed the quality control. RNA samples (derived from blood) were processed for RNA Seq with polyA selection and sequenced on Illumina HiSeq High Output mode with a sequencing configuration of 2×150 paired-end reads (GENEWIZ, South Plainfield, NJ). A total of 10M paired reads per sample was set as a threshold to account for high globin reads; 29 samples were re-sequenced to meet threshold.

### Gene expression quality control analysis

We processed whole-blood gene expression data using the RAPiD.19 RNA-sequencing pipeline, and calculated normalized TPM counts from RSEM.^26^ We performed quality control analysis on the counts to verify sequencing and residual contributions to variance using VariancePartition^27^. We corrected each sequencing batch for sex, age, and genotype-derived principal components using Limma/voom weighted least-squares linear regression.^28^ We rank normalized and combined the residuals from the linear regression of each batch, and these values were used in all subsequent association tests for CAPS total score and five PTSD dimension scores (re-experiencing, avoidance, numbing, dysphoric arousal and anxious arousal).

### Differential gene expression analysis

We used whole blood RNA-sequencing to test for associations between gene expression and WTC-related PTSD symptom severity on (i) highest lifetime CAPS score (CAPS_L_), (ii) past-month CAPS score (CAPS_PM_), and (iii) PTSD dimension scores including re-experiencing, avoidance, numbing, dysphoric arousal and anxious arousal, correcting for batch and surrogate variables (Eqn. 1).

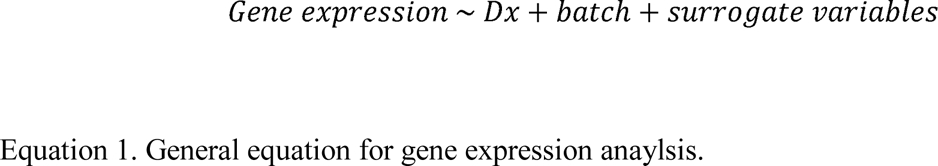

In addition, study participants had a wide range of psychiatric and somatic comorbidities, including some with substantial shared genetic aetiology and overlapping symptom profiles (e.g., major depressive disorder); comorbidities that may represent systemic manifestations of of PTSD (e.g., cardiovascular disease^29^); and exposure to the dust cloud during and following 9/11. We expect all of these factors to have substantial impacts on gene expression. Significant co-linearity between some of these measures and CAPS scores preclude including these variables as covariates within our analysis, and testing directly for their effect on gene expression (in particular, due to high correlations between length of time at the WTC site, CAPS score, and dust-cloud exposure; and between CAPS score and co-morbid medical disorders potentially constituting systemic manifestations of PTSD). Instead, in order to test whether these comorbidities and exposures might account for some of the CAPS-expression associations we observe, we also tested for gene expression associations with (i) an index of dust cloud exposure^30^; and (ii) number of medical comorbidities. Next, we tested for (i) interaction effects between each of these measures and CAPS score; (ii) enrichment of genome-wide significant associations between these measures and CAPS score; and (iii) genome-wide correlations in association statistics. For all gene expression analyses, we established significance using a Benjamini-Hochberg^31^ FDR correction < 5%.

### Gene-set enrichment of PTSD-associated genes

We tested for gene set enrichment among our genes associated with CAPS_L_, CAPS_PM_, and PTSD dimension scores by (i) analyzing the significant genes from the association tests for pathway enrichment by gene permutation testing and (ii) analyzing all genes from the ranked association test gene lists to subject permutation using the R versions of GSEA^32^ and fgsea^33, 34^. For gene permutation testing, we included all nominally significant genes (p<0.05), and tested for association with 105 gene sets using Kyoto Encyclopedia of Genes and Genomes (KEGG) database^35^ for pathway enrichment.

We applied phenotype permutation testing rather than gene set permutation to keep the correlations between the genes in the dataset and the genes in the gene set pathways. For the subject permutation testing, each test was run with 10000 permutations, and pathways that passed Benjamini-Hochberg multiple testing correction were considered significantly enriched. To synthesize this large amount of gene set information, we generated comparative PTSD symptom plots using Clusterprofiler^36^ in R. Comparative gene set plots contained pathways which passed FDR<0.05 significance threshold.

### Meta-analysis with existing gene expression analyses

To replicate our gene expression results, we meta-analyzed our data with association statistics from five other genome-wide gene expression analyses:

1. **WTC responder study Stony Brook University (SB WTC)** ^3^; N=282. Data are divided into discovery (WTC-d) and replication (WTC-r) cohorts.
2. **Trauma Mega-Analysis study (TMA)** ^12^. TMA combines 7 different PTSD studies and transformed the expression data into three categories: combat (N=169), male interpersonal (N=112), and female interpersonal trauma (N=259)^12^. We analyzed these data as separate trauma studies for the purposes of our meta-analysis (Table 2).

**Table 2.** Medical comorbidities and corresponding N of World Trade Center first responders with each comorbidity.

| **Trauma Study** | **ISMMS** | **WTC-d<sup>3</sup>** | **WTC-r<sup>3</sup>** | **TMA-C<sup>12</sup>** | **TMA-MI<sup>12</sup>** | **TMA-FI<sup>12</sup>** |
| --- | --- | --- | --- | --- | --- | --- |
| Past-month PTSD/control, N subjects | 53/302 | 57/138 | 24/63 | N/A | N/A | N/A |
| Lifetime PTSD/Control, N subjects | 108/247 | N/A | N/A | 85/84 | 45/67 | 99/160 |
| Total N subjects | 355 | 195 | 87 | 169 | 112 | 259 |
| Available genes (p<0.05), N | 1016 | 27 | 27 | 806 | 418 | 418 |
| Age, Mean | 54.1 (8.3) | 52 (8.12) | 52 (8.12) | 24.4 (4.7) | 41.1 (12.8) | 39.5 (12.3) |
| Male, N (%) | 291 (82) | 195 (100) | 87 (100) | 169 (100) | 112 (100) | 0 (0) |
| European American, N (%) | 216 (60.8) | 165 (84.6) | 74 (84.6) | 108 (63.6) | 34 (30.3) | 62 (23.9) |
| African American, N (%) | 64 (18) | N/A | N/A | N/A | 78 (69.7) | 197 (76.1) |
| Hispanic, N (%) | 72 (20.3) | N/A | N/A | N/A | N/A | N/A |

Note: ISMMS=Icahn School of Medicine at Mount Sinai (this study); WTC-d=Stony Brook World Trade Center discovery cohort; WTC-r=Stony Brook World Trade Center replication cohort; TMA-C=Trauma mega-analysis combat cohort; TMA-MI=Trauma mega-analysis male-interpersonal cohort; TMA-FI=Trauma mega-analysis female-interpersonal cohort. <sup>3</sup>Kuan PF et al 2017,
<sup>12</sup>Breen MS et al 2018.

Since the majority of studies focus on PTSD case/control status, rather than associations with continuous CAPS scores (as here), we repeated our analysis to compare gene expression between PTSD cases (defined as meeting DSM-IV criteria for PTSD and a total CAPS ≥ 40) and controls (all others in our sample).

We meta-analyzed PTSD case-control association statistics using a sample-size based meta-analysis approach in METAL^37^. We included all genes from our analysis that reached p<0.05 (N=9,380 for past-month and N=1,016 for lifetime) and all available genes from other studies (N=27-806; Table 2).

### Cellular deconvolution associated with CAPS scores

We applied CIBERSORT to our raw counts matrix to deconvolute immune cell types in our patients using the immune cell matrix reference panel LM22^38^. We tested for association between cell type proportions and CAPS_PM,_ CAPS_L_.

## Results

### Gene expression is associated with PTSD symptom levels in WTC first responders

We tested for association between expression of 12,220 genes and total CAPS score in a sample of 355 WTC first responders. We identified 31 genes significantly associated with total past-month CAPS score (CAPS_PM_; Figure 3), and 66 genes associated with lifetime (highest) CAPS score (CAPS_L_, Figure 3). Of these, 42/66 genes are associated only with CAPS_L_, (and not CAPS_PM_), while 7/31 genes were associated only with CAPS_PM_, (and not CAPS_L_). Genome-wide associations with CAPS_PM_ and CAPS_L_ were significantly correlated (Beta ρ = 0.82, p<2.2×10^-16^; FDR-adjusted P-values ρ = 0.79, p<2.2×10^-16^) (Table 3).

**Figure 3.**
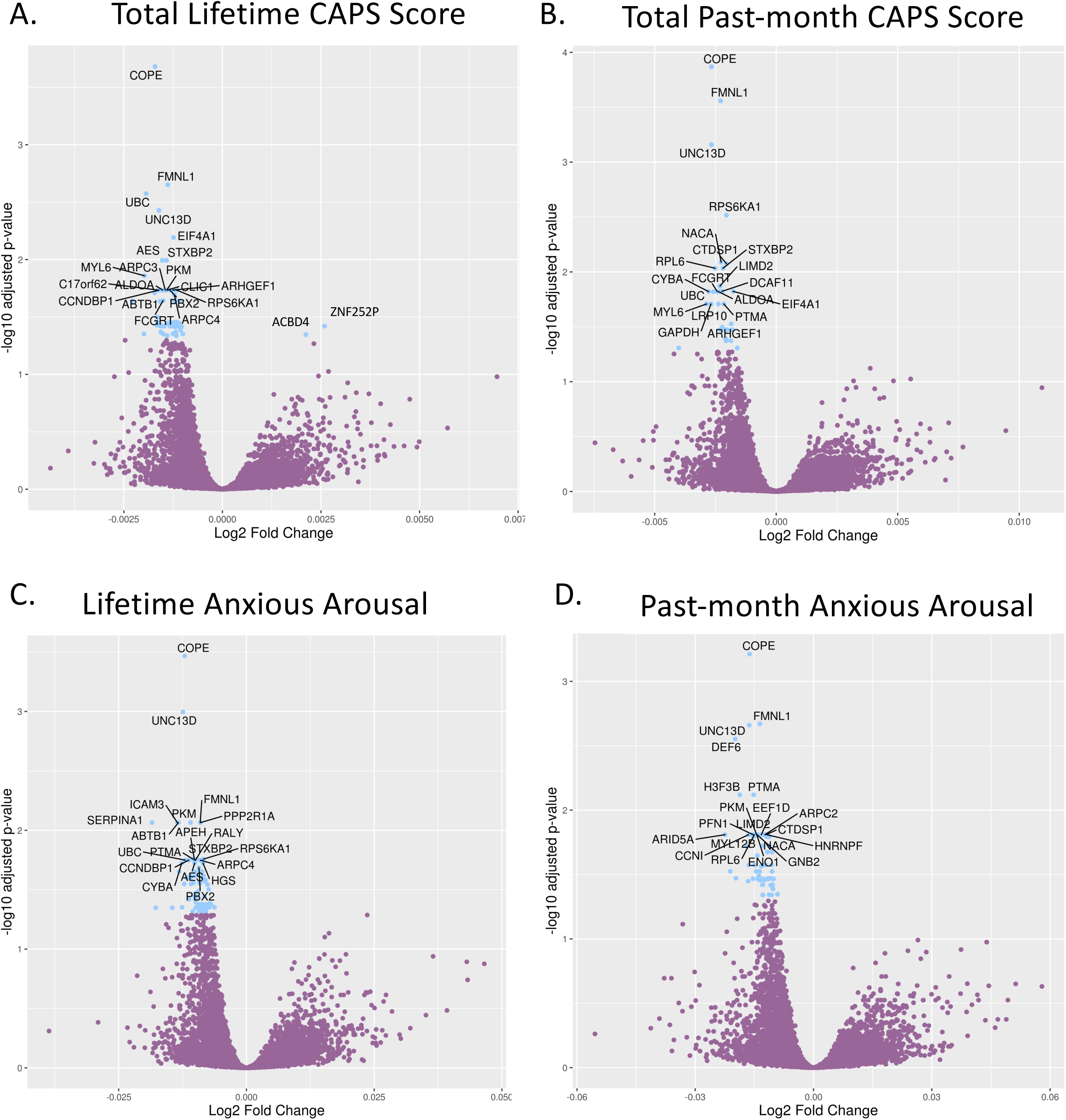
**Differential gene expression analysis** of A) lifetime CAPS B) CAPS past-month C) anxious arousal lifetime and d) anxious arousal past-month in N=355 World Trade Center first-responders. Total number of differentially expressed genes (p<0.05): A) N=66, B) N=31, C) N=86, D) N=61. Phenotypes were corrected for sex, age, batch, and first 10 ancestry principal components.

**Table 3.** Significant (p<0.05) genes associated with each phenotype and corresponding statistical values: log2 fold change, average expression, t-statistic, p value, FDR-corrected p value, and beta value. Phenotypes included are lifetime CAPS and past-month CAPS and all symptom dimensions.

| **Lifetime vs. Past-month** | **R<sup>2</sup> (of beta values)** | **p** | **R<sup>2</sup> (of FDR-adjusted P values)** | **p** |
| --- | --- | --- | --- | --- |
| Total CAPS score | 0.82 | <2.2x10<sup>-16</sup> | 0.79 | <2.2x10<sup>-16</sup> |
| Re-experiencing | 0.66 | <2.2x10<sup>-16</sup> | 0.63 | <2.2x10<sup>-16</sup> |
| Anxious Arousal | 0.8 | <2.2x10<sup>-16</sup> | 0.77 | <2.2x10<sup>-16</sup> |
| Avoidance | 0.73 | <2.2x10<sup>-16</sup> | 0.71 | <2.2x10<sup>-16</sup> |
| Dysphoric Arousal | 0.75 | <2.2x10<sup>-16</sup> | 0.72 | <2.2x10<sup>-16</sup> |
| Numbing | 0.66 | <2.2x10<sup>-16</sup> | 0.64 | <2.2x10<sup>-16</sup> |

**Bivariate Correlations Between PTSD Symptom Dimensions**
| **Past-month** | Re-experiencing | Anxious Arousal | Avoidance | Dysphoric Arousal | Numbing |
| --- | --- | --- | --- | --- | --- |
| Re-experiencing | 1 | 0.53 | 0.63 | 0.59 | 0.62 |
| Anxious Arousal |  | 1 | 0.57 | 0.64 | 0.55 |
| Avoidance |  |  | 1 | 0.61 | 0.63 |
| Dysphoric Arousal |  |  |  | 1 | 0.71 |
| Numbing |  |  |  |  | 1 |
| **Lifetime** | Re-experiencing | Anxious Arousal | Avoidance | Dysphoric Arousal | Numbing |
| Re-experiencing | 1 | 0.72 | 0.79 | 0.75 | 0.73 |
| Anxious Arousal |  | 1 | 0.71 | 0.73 | 0.69 |
| Avoidance |  |  | 1 | 0.74 | 0.79 |
| Dysphoric Arousal |  |  |  | 1 | 0.78 |
| Numbing |  |  |  |  | 1 |

**Note:** CAPS = Clinician-Administered PTSD Scale; FDR = false discovery rate.

We tested for enrichment of 59 well-studied PTSD candidate genes^11^ (Sup. Table 1) within our association statistics. We did not observe enrichment of these genes within our past-month or lifetime association analyses (p=0.174, 0.245). However, of these candidate genes, *SERPINA1* was significantly associated with CAPS_L_ score.

### Environmental exposure to the WTC dust cloud

Next, we tested whether our genes associated with CAPS scores are specific to PTSD, or are driven by spurious associations with comorbid diagnoses or environmental exposure to the dust cloud at Ground Zero. In particular, a number of individuals within our study have comorbid conditions and complex medical histories (Table 4), including disorders with substantial shared genetic and environmental etiology with PTSD, and disorders and traits that may present as systemic manifestations of PTSD.

**Table 4.** PTSD and demographic information of meta-analysis study. Includes Stony Brook World Trade Center cohort and a mega-analysis study split into combat, male interpersonal and female interpersonal trauma.

| **Conditions** | **N (%)** |
| --- | --- |
| Hypertension | 162 (45.6) |
| Cardiac disease | 28 (7.9) |
| Diabetes | 44 (12.4) |
| GERD | 184 (51.8) |
| Chronic pain | 144 (40.6) |
| Arthritis | 138 (38.9) |
| Asthma | 161 (45.4) |
| Cancer | 51 (14.4) |
| Chronic rhinitis | 169 (47.6) |
| Sleep apnea | 127 (35.8) |
| Kidney disease | 8 (2.3) |
| Chronic skin condition | 70 (19.7) |
| High cholesterol | 174 (49) |
| Liver disease | 15 (4.3) |
| Migraine | 50 (14.1) |
| Osteoporosis | 11 (3.1) |
| Rheumatoid Arthritis | 15 (4.3) |
| Stroke | 4 (1.1) |
| Traumatic brain injury | 5 (1.4) |

Note: GERD = gastroesophageal reflux disease.

First, we tested whether these medical comorbidities alone may account for the associations we observe; we identified 175 gene associations with an aggregate summary score of medical comorbidities. Notably, these genes do not include any of our significant associations with CAPS scores. We identified only one gene, *STX10,* with significant interaction between CAPS scores and comorbid conditions; that is, gene expression was elevated specifically among individuals with both high CAPS_L_ and a large number of comorbid diagnoses (Figure 4).

**Figure 4.**
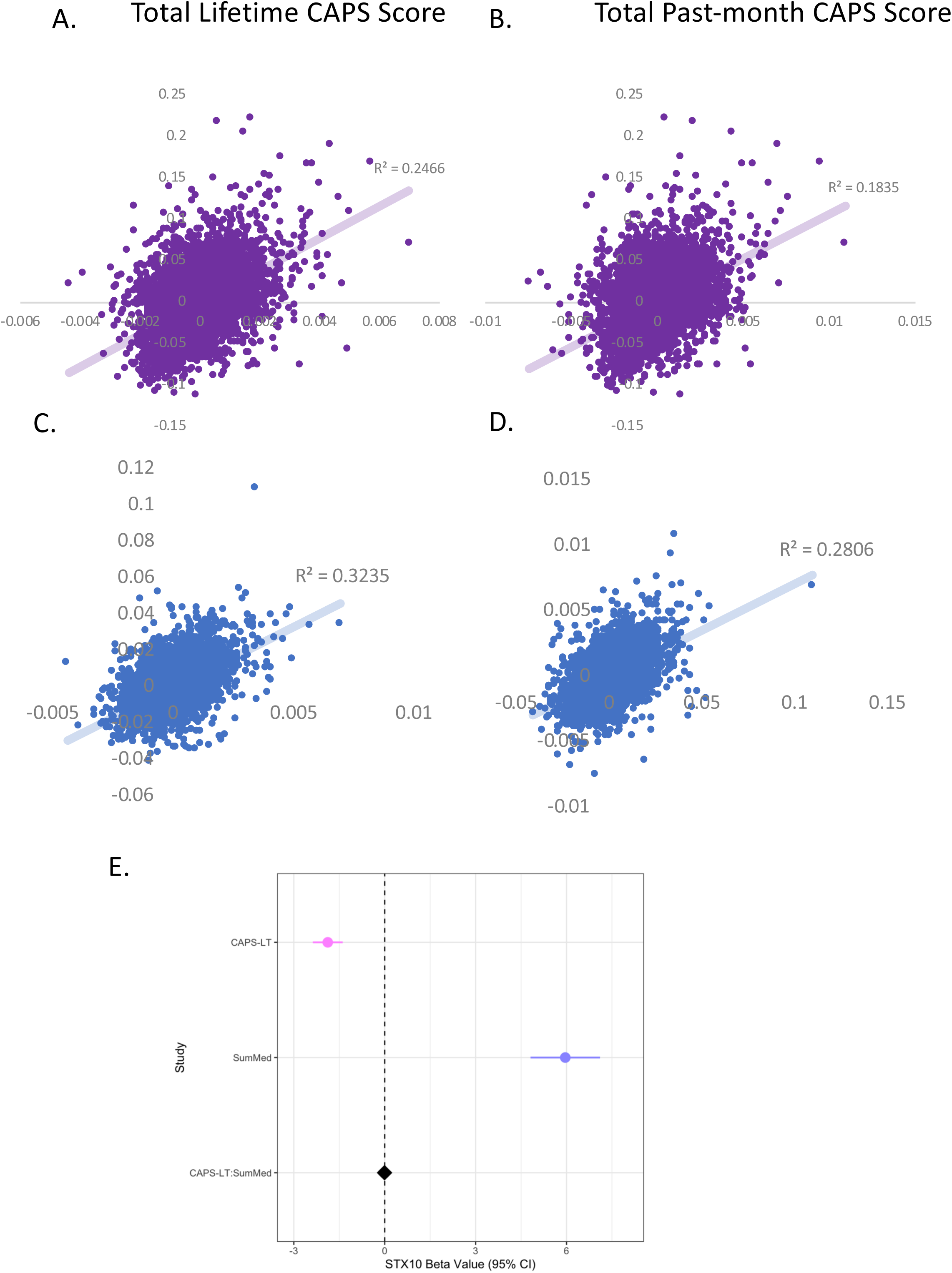
**Dust cloud severity (A-B) and disease comorbidity (C-D) of CAPS lifetime and past-month genome-wide correlations. (E) Interaction term, *STX10*, of disease comorbidity and CAPS lifetime.** SumMed: summary of medical terms for disease comorbidity; CAPS-LT: CAPS lifetime.

Next, we tested for association between gene expression and exposure to the dust cloud at Ground Zero^30^. We identified 561 genes significantly associated with this exposure. We tested for, but did not find, any significant interactions between CAPS scores and dust cloud exposure (p>0.05), and did not observe a large correlation of expression results in genome-wide significant genes between the two analyses for CAPS_PM_ or CAPS_L_ (R^2^=0.1835, R^2^=0.2466, p<0.05) (Figure 4). Together, these analyses imply that our gene-CAPS score associations are specific and relevant to PTSD, rather than due to confounding by comorbid diagnoses or dust cloud exposure.

### Gene expression analysis reveals PTSD dimension-specific associations

Next, we tested for gene expression associations with past-month and lifetime PTSD symptom dimensions (re-experiencing, avoidance, dysphoric arousal, anxious arousal, numbing; Sup. Table 2).

Our analysis revealed overlapping and unique genes for each symptom dimension, and significant correlation of genome-wide association statistics between symptom dimensions. In particular, both our past-month and lifetime analyses identified a large number of genes (61 and 82, respectively; Sup. Table 2) associated with anxious arousal, including many genes not associated with any other phenotype in our analysis (16, 27, respectively). By contrast, although we identified a substantial number of genes significantly associated with dysphoric arousal (20, 16 for past-month and lifetime respectively), only two genes were uniquely associated with this phenotype (1 gene in past-month; 2 in lifetime). Only one gene, *COPE*, was significantly associated with every phenotype tested in our analysis. A second gene, *EIF4A1,* was also significantly associated with every lifetime (highest) phenotype (Figure 5).

**Figure 5.**
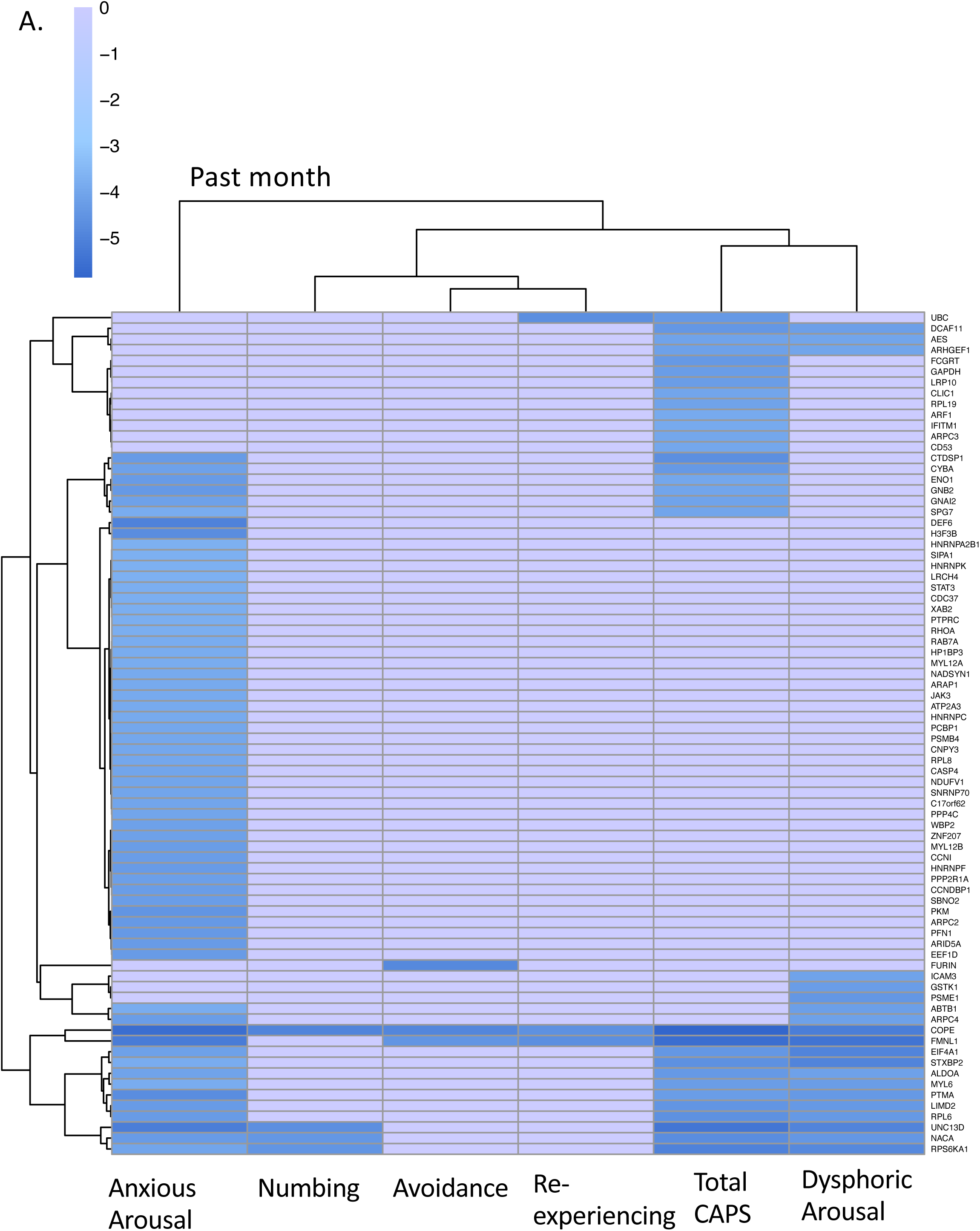
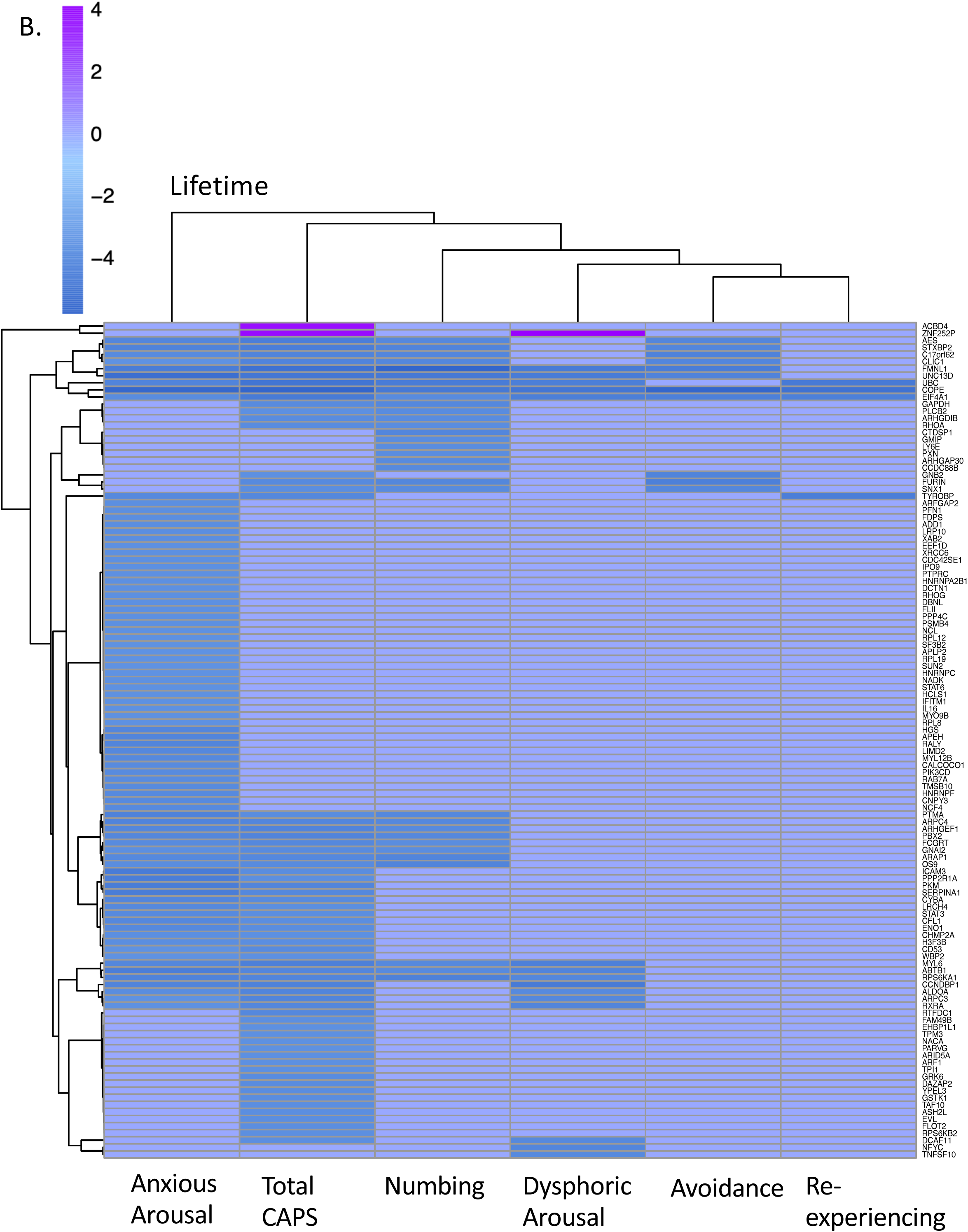
**Heatmap of differentially expressed (p<0.05) genes** from a gene expression analysis across (A) past-month CAPS and symptom dimensions and (B) lifetime CAPS and symptom dimensions in N=355 World Trade Center first-responders. Phenotypes were corrected for sex, age, batch, and first 10 ancestry principal components. Gene clusters represent genealogical expression diversity among lifetime total CAPS and symptom dimensions.

### PTSD pathway enrichment demonstrates immune, psychiatric, and metabolic relationships

Our genetic enrichment and pathway analyses identified well-established PTSD mechanisms and pathways, including pathways associated with inflammation, neurological signaling pathways, structural remodeling within and between cells, and HPA-axis and signaling^39–44^. Based on our KEGG pathway enrichment analysis, we revealed a set of genes significantly associated with psychiatric disorders pathways that were significantly enriched in our results: *FURIN, PPP2R1A, GNAI2, PCLB2,* and *GNB2.* The two symptoms that we discovered were associated with *FURIN* and their FDR-corrected p-values were avoidance (p=0.028, B=2.55) and numbing (p=0.034, B=-0.009). *PPP2R1A* was associated with anxious arousal (p=0.009, B=2.60). *PCLB2* was only associated with numbing (p=0.046, B=-0.952). *GNAI2* was associated with anxious arousal (p=0.042, B=-0.780) and numbing (p=0.047, B=-1.03), and *GNB2* was only associated with avoidance (p=0.036, B=1.130) (Figure 6).

**Figure 6.**
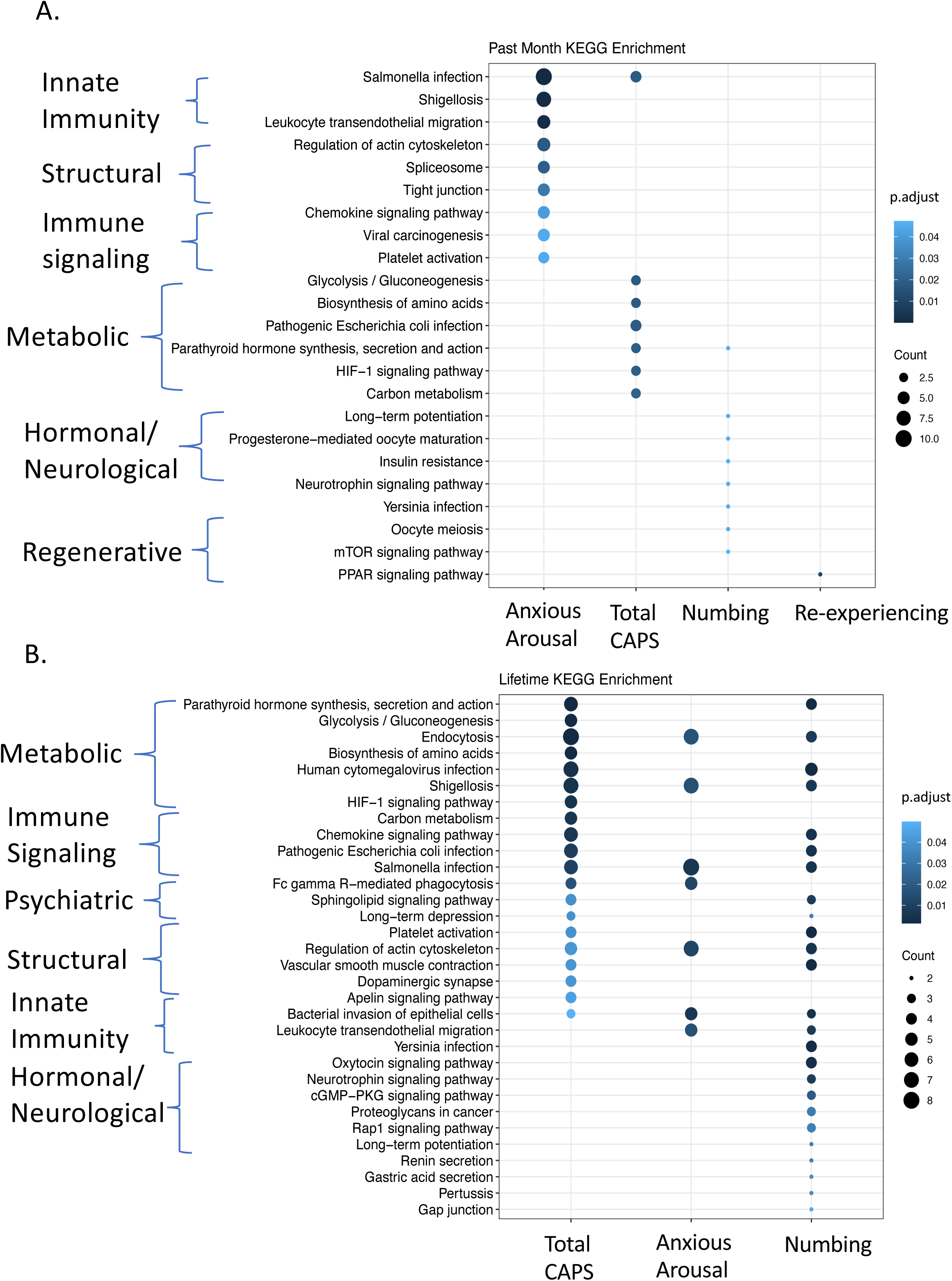
**KEGG pathway enrichment** of differentially expressed genes in N=355 World Trade Center first-responders in (A) past-month and (B) lifetime CAPS. Pathways categorically grouped by metabolic, immune signaling, innate immunity, structural, hormonal/neurological, regenerative and psychiatric function.

Immunological and metabolic gene enrichment was consistent across CAPS_L_ score and numbing, but was less pronounced in anxious arousal. For past-month analyses, immune function was most associated with anxious arousal, whereas metabolic function was enriched in CAPS_PM,_ and numbing, to a lesser extent. For both lifetime and past-month scores, neurological signaling pathways were most significantly pronounced in numbing, and were less prevalent in the overall total CAPS score analysis. For lifetime scores, structural pathway enrichment was significantly higher in total CAPS score, anxious arousal, and numbing, whereas for past-month scores, structural enrichment was most associated with anxious arousal (Figure 6).

### Meta-analysis prioritizes 10 genes associated with PTSD

We sought to replicate our associations with previous PTSD studies. Since the majority of publicly available PTSD gene expression analyses follow a case-control, rather than quantitative measure (CAPS score) analysis, we converted our continuous CAPS_PM_ and CAPS_L_ values to PTSD case/control using (CAPS≥ 40) and DSM-IV PTSD-criteria, and repeated our analysis. Our case/control and CAPS score association statistics were significantly correlated (ρ=0.72, p<2.2×10^-16^, ρ=0.79, p<2.2×10^-16^); however, we note a substantial decrease in the number of significantly associated genes when using a case-control design, compared to our initial quantitative analysis, as we would expect^16^. Twelve genes were significant for past-month PTSD (PTSD_PM_) case/control and 22 genes were significant for lifetime PTSD (PTSD_L_) case/control, versus 31 genes for CAPS_PM_ and 66 genes for CAPS_L_.

We meta-analyzed our results with five publicly available cohorts (N=739 cases, 438 controls); two including WTC responders, and three including combat and interpersonal trauma (Table 2). For PTSD_PM_ we identified 5 significant genes *–COPE, CIRBP, FCGRT, NACA,* and *ZNF429* (p<5.33×10^-6^)*–*, and for PTSD_L_ 8 significant genes *–COPE*, *CIRBP, TMSB10, FCGRT, CLIC1, RPS6KB2, HNRNPUL1* and *ALDOA* (p<4.92×10^-5^)*–*, including genes associated with inflammation and immune response (Figure 7). Of these 10 genes, only one (*COPE*) had significant heterogeneity of effect size between cohorts: our study and TMA-combat. Three further genes were unique to our study; *NACA* p=3.34×10^-6^, *CLIC1,* p=1.9×10^-5^, and *HNRNPUL1* p=4.08 x10^-5^ (Figure 7). The remaining six genes were significant across multiple studies in our meta-analysis, with highly consistent (negative) direction of effect (i.e., consistently decreased expression in cases compared to controls)*: ZNF439* (p=4.78×10^-6^), *CIRBP* (p=1.29 x10^-6^), *TMSB10* (p=6.31 x10^-6^), *FCGRT* (p=1.12 x10^-5^), *RPS6KB2* (p=3.47 x10^-5^), and *ALDOA* (p=4.66 x10^-5^) (Table 5).

**Figure 7.**
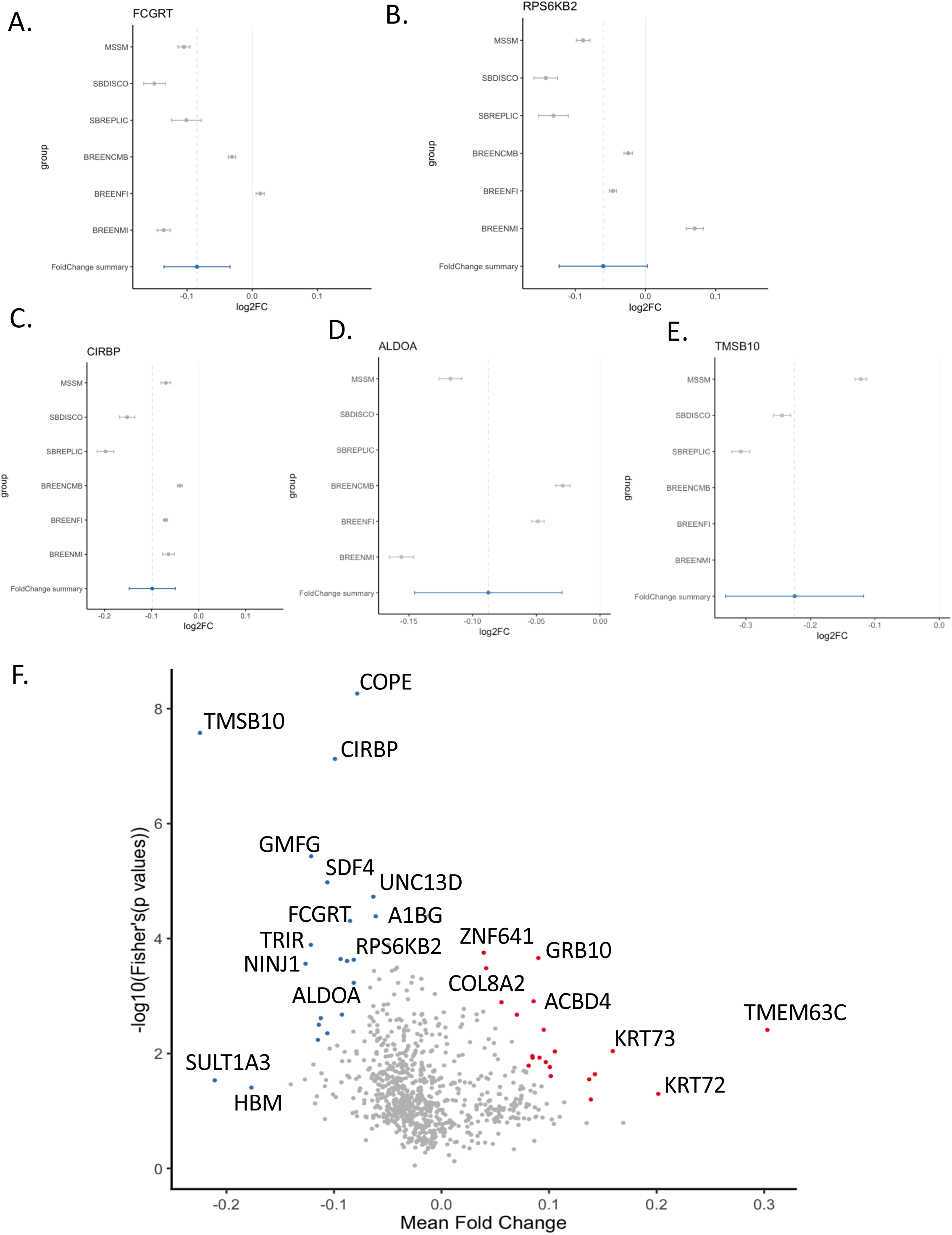
Forestplots (A-E) and volcano plot (F) of genome-wide significant genes from our meta-analysis. Meta-analysis included our gene expression analysis, another World Trade Center (WTC) study, and a mega-analysis of 7 different compiled trauma studies.

**Table 5.** Table of genome-wide significant genes from our meta-analysis for lifetime and past-month PTSD. Meta-analysis included our gene expression analysis, another World Trade Center (WTC) study, and a mega-analysis of 7 different compiled trauma studies. Direction is defined as our study, WTC discovery cohort, WTC replication cohort, military trauma, male interpersonal trauma and female interpersonal trauma (from the mega-analysis study).

| Marker Name | Weight | Z-score | P-value | Direction | Heterogenous I-Squared | Heterogenous Chi-Squared | Heterogenous degrees of freedom | Heterogenous P value |
| --- | --- | --- | --- | --- | --- | --- | --- | --- |
| COPE | 524 | -5.277 | 1.31E-07 | -??-?? | 91.7 | 12.103 | 1 | 0.0005034 |
| CIRBP | 982 | -4.842 | 1.29E-06 | --?--- | 0 | 3.785 | 4 | 0.4358 |
| TMSB10 | 442 | -4.516 | 6.31E-06 | --???? | 32 | 1.47 | 1 | 0.2253 |
| FCGRT | 1177 | -4.392 | 1.12E-05 | ----+- | 52 | 10.423 | 5 | 0.06411 |
| ALDOA | 895 | -4.072 | 4.66E-05 | -??--- | 43.2 | 5.281 | 3 | 0.1523 |
| RPS6KB2 | 1177 | -4.14 | 3.47E-05 | -----+ | 50.5 | 10.099 | 5 | 0.07248 |
| HNRNPUL1 | 355 | -4.103 | 4.08E-05 | -????? | 0 | 0 | 0 | 1 |
| CLIC1 | 355 | -4.276 | 1.90E-05 | -????? | 0 | 0 | 0 | 1 |

**Past-month PTSD**
| Marker Name | Weight | Z-score | P-value | Direction | Heterogenous I-Squared | Heterogenous Chi-Squared | Heterogenous degrees of freedom | Heterogenous P value |
| --- | --- | --- | --- | --- | --- | --- | --- | --- |
| COPE | 524 | -4.778 | 1.77E-06 | -??-?? | 89.8 | 9.826 | 1 | 0.00172 |
| CIRBP | 982 | -5.016 | 5.27E-07 | --?--- | 0 | 3.676 | 4 | 0.4516 |
| FCGRT | 1177 | -4.793 | 1.64E-06 | -----+ | 59.9 | 12.475 | 5 | 0.02883 |
| ZNF439 | 451 | -4.574 | 4.78E-06 | ?---?? | 0 | 1.695 | 2 | 0.4285 |
| NACA | 355 | -4.649 | 3.34E-06 | -????? | 0 | 0 | 0 | 1 |

Note: CAPS-5 = Clinician-Administered PTSD Scale. Direction is defined as our study, WTC discovery cohort<sup>3</sup>, WTC replication cohort<sup>3</sup>, military trauma, male interpersonal trauma and female interpersonal trauma (from mega-analysis study<sup>12</sup>). <sup>3</sup>Kuan PF et al 2017, <sup>12</sup>Breen MS et al 2018.

### Cellular deconvolution identifies differences in cell populations between responders with PTSD and controls

Since many of our PTSD-associated genes are related to immune function, we tested whether immune cell type proportions were correlated with CAPS scores in individuals in our sample. We performed cell-type deconvolution to identify cell-type proportions for 22 cell types across all 355 individuals in our sample. We found significant increase of CD4 naïve T cell (p<0.0049) proportions with CAPS_PM_, and significant increase of eosinophils (p< 0.042) and CD4 memory resting T cells (p<0.044) associated with CAPS_L_. In addition, we found significant decrease of activated natural killer cells (p<0.040) associated with CAPS_L._ (Figure 8).

**Figure 8.**
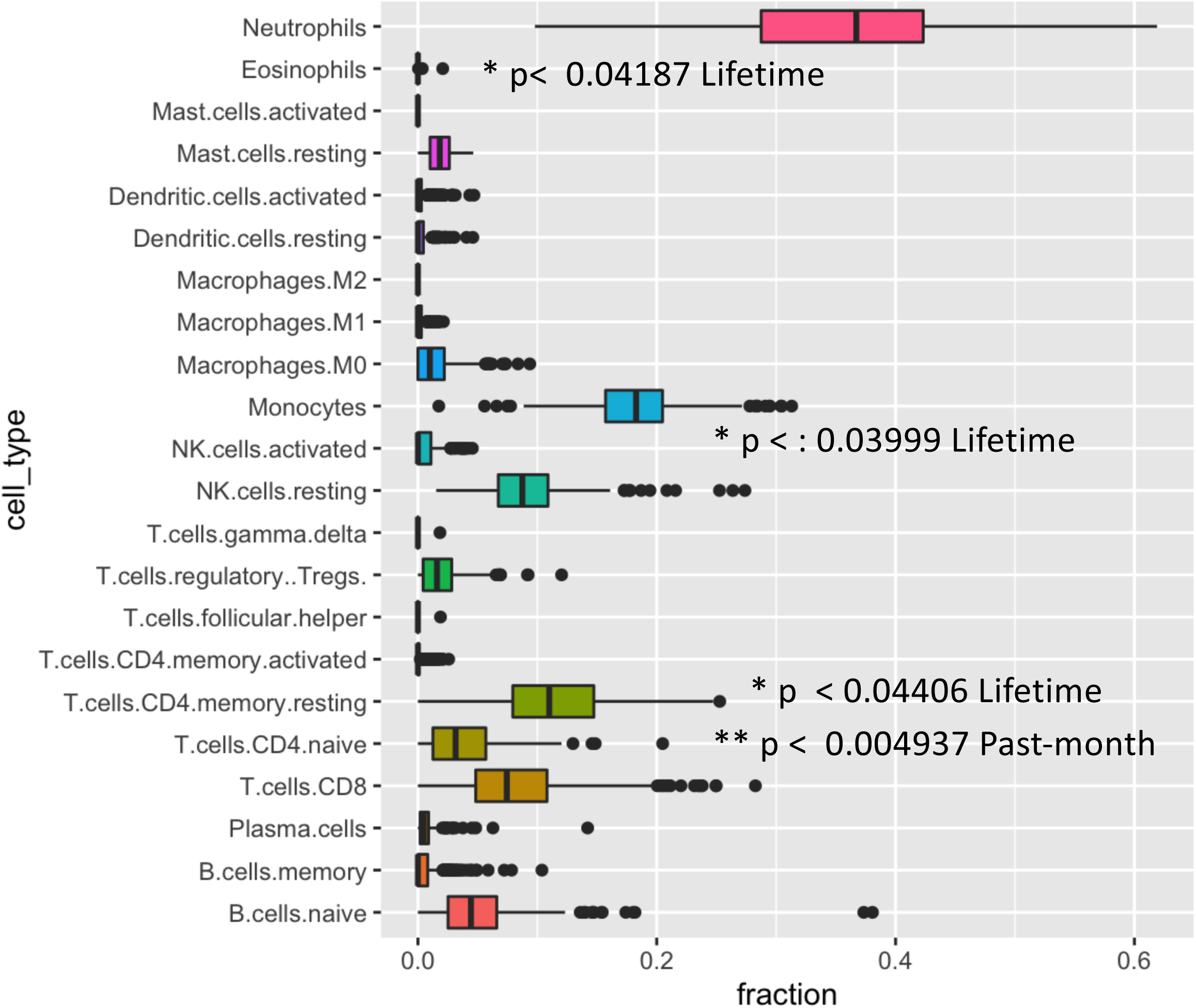
Cellular deconvolution of immune cells in peripheral blood. Significant cell type differences for CAPS lifetime and past-month compared to controls are noted.

## Discussion

Although trauma is ubiquitous as a human experience, the types of traumatic experiences vary greatly among individuals. Our study sample and design present a unique opportunity to examine gene expression and PTSD symptoms related to a particular traumatic experience, the 9/11 WTC disaster and its aftermath. To our knowledge, ours is the largest single-traumatic event gene expression dataset to date. Using CAPS score as a quantitative measure of WTC-related PTSD symptom severity, we found 66 genes associated with lifetime (highest) CAPS score and 31 genes associated with past-month CAPS score. We additionally found interaction between CAPS_L_ and one gene, *STX10*, in medical comorbidities. Pathway analysis links our associated genes to metabolic, immunological, structural, and neurological pathways. In addition, we found 10 genes associated with lifetime and past-month PTSD: *COPE, CIRBP,* and *FCGRT* shared between the two; *NACA* and *ZNF429* specific to past-month PTSD; and *TMSB10*, *CLIC1, RPS6KB2, HNRNPUL1* and *ALDOA* specific to lifetime PTSD. Furthermore, cellular analysis of our cohort demonstrated an enrichment in CD4 T cells and eosinophils in responders with PTSD. Additionally, natural killer cells were decreased in these patients. Our findings support previous studies which tie together the immune system and PTSD as a systemic disease. Ultimately, these results represent an additional wealth of knowledge to help understand the genetic expression and biological etiology of PTSD and uncover potential biomarkers for the disease.

Examining both lifetime and past-month CAPS scores allowed us to look at not only chronic effects of PTSD but longevity of the outcomes. While CAPS_PM_ is considered the standard for case/control analysis, looking at both CAPS_L_ and CAPS_PM_ allows us to ask more specific questions about the role and relevance of elevated gene expression in PTSD. For example, genes associated with CAPS_PM_ might represent current expression changes in PTSD, while those associated with CAPS_L_ (lifetime CAPS score representing, for each responder, the highest WTC-related PTSD symptom levels ever reached since 9/11/2011) may represent long-lasting expression changes resulting from lifetime PTSD.

We identified genes associated with past-month and lifetime CAPS scores that have been associated with other psychiatric disorders, such as major depressive disorder, schizophrenia and autism: *FURIN, PPP2R1A, PLCB2, GNAI2, and GNB2* (Figure 6)^45–57^. We additionally identified a group of genes as significantly associated with several lifetime and past-month PTSD symptom dimensions (*SERPINA1, RPS6KA1,* and *STAT3*) that have been previously linked to PTSD pathophysiology^1, 27–31^, and genes associated specifically with anxious arousal that have been previously associated with anxiety disorders: *DCTN1,* and *FLI1*^58, 59^. One gene in our study, *COPE*, was associated with every PTSD phenotype in this analysis. Until now, *COPE* has not been well studied in the context of psychiatric disorders^60^, but it has been implicated in the context of Alzheimer’s disease^61, 62^. Canonically, the COPE protein is the epsilon subunit of the coatomer protein complex I (COPI) which regulates endocytosis from the plasma membrane and is involved in Golgi-to-lysosome transportation. Other subunits of COPI have been implicated in hereditary diseases that cause microcephaly and in autoimmune disorders^63–65^.

Importantly, our analytical design allowed us to test for potential confounding effects of comorbidities and environmental exposures^66, 67^. We expected many of these comorbidities and exposures to have a significant impact on gene expression, particularly as some comorbidities may be more recently occurring than, for example, the highest CAPS_L_ measure. Therefore, we tested directly for genes associated with (i) comorbidities and (ii) dust cloud exposure. Although we identified a large number of genes associated with each phenotype, we note that our associations do not overlap with our top PTSD genes; we did not observe significant enrichment of shared associations, and only observed one gene with significant interaction effect between comorbidities and CAPS_L_. Therefore, we conclude that our results are not confounded by these exposures; our gene associations are specific to PTSD rather than broadly corresponding to exposure or general ill-health.

KEGG enrichment of our associated genes determined genetic changes in metabolic, immunological, structural, and neurological pathways associated with total CAPS, numbing, and anxious arousal phenotypes (Figure 7). Anxious arousal and numbing tend to have few significantly associated genes in common, while both past-month and lifetime CAPS scores display a few unique genes but many shared ones. *GNAI2*, while associated with many numbing-related pathways (long-term depression, oxytocin signaling, etc.), is also significantly associated with some peripheral anxious arousal-related and CAPS_L_–related pathways (leukocyte migration, and chemokine signaling, respectively). On the other hand, anxious arousal is uniquely associated with genes such as *HGS, ARFGAP2* and *RAB7A*, which are linked to endocytosis and immunity. Similarly, our past-month and lifetime CAPS phenotypes are uniquely associated with glycolysis/gluconeogenesis for genes *ALDOA, GAPDH, ENO1, TPI1,* and *PKM*, which may play roles in HPA-axis or metabolic dysregulation (Table 4).

Our analyses identified 10 genes reaching genome-wide significance. Of these, 3/10 (*NACA* p=3.34×10^-6^, *CLIC1,* p=1.9×10^-5^, and *HNRNPUL1* p=4.08 x10^-5^) are specific to our study; all three are downregulated in WTC responders with PTSD compared to controls. *NACA* has been previously studied as a potential biomarker for depression in mice under stress conditions^68^. The function of *HNRNPUL1* is relatively unknown. Studies have suggested it might play a role in DNA damage repair and nucleocytoplasmic RNA transport. *CLIC1* has been implicated in other psychiatric disorders, but its primary function is inflammasome regulation^69–71^. In addition, 6/10 genes had highly consistent directions of effect. These include *FCGRT* (p=1.12x^-05^), which has been characterized as an immune regulator of dendritic cell cross-presentation of IgG immune complexes, necessary to activate a cytotoxic T-cell response and clear antigens^72^. Our analysis demonstrates a down-regulation of *FCGRT* in WTC responders with PTSD, suggesting reduced IgG immune complex clearance in these patients (Figure 6).

There is evidence that macro-and micro-level physiological damage is a fundamental component of PTSD, as well as cytoskeletal restructuring for fear-based memory formation in the amygdala^43, 44^. In this study we observed decreased expression in *CIRBP* (p= 1.29x^-06^), a protein that traditionally regulates stress and apoptosis under conditions of extreme cold (Figure 6). Its role as a potential biomarker has been previously explored for different psychiatric disorders^73, 74^. The decrease observed in *TMSB10* expression also contributes to the dysregulation of apoptosis. *TMSB10* is a pro-apoptotic protein that has been previously associated with downregulation of gene expression after trauma exposure^3, 75, 76^. In our meta-analysis, the directionality of effect (p<2.2x^-16^) for each gene was decreased, consistent with PTSD pathophysiology^77, 78^.

Since many of our PTSD-associated genes are related to immune function, we tested for the enrichment of immune cell types in our study. We found an overall enrichment of CD4-positive T cells for both past-month and lifetime CAPS scores (Figure 7), consistent with previous studies^79, 80^, including among WTC responders^8^. In addition to CD4 T cell enrichment, our study also found enrichment in eosinophils and a decrease in natural killer cells for CAPS_L_ (Figure 7). These cellular diversities may point to a higher inflammatory signature in PTSD, particularly in the case of CD4 T cell enrichment. It has been well demonstrated that dysregulation of CD4 T cells leads to autoimmune activation^81^, and in combination with an increase in eosinophils can lead to an inflammatory cascade in patients. There is strong evidence that PTSD is associated with a pro-inflammatory state, which our findings support^82–86^.

While our study provides an in-depth look at the genetic expression and outcomes related to a specific traumatic experience, we note some significant caveats. Our expression analysis was limited to blood, but should be expanded to other tissues in the future, such as the brain. Similarly, our analysis was restricted to whole blood, but a more in-depth single cell analysis will be critical to determine gene expression in individual cell types. In addition, we note that our cohort includes a significant proportion of individuals who have self-selected into high-risk professions. As such, we expect a higher lifetime exposure to stressful situations, including potentially many other life-threatening scenarios. It is likely that the PTSD symptoms observed here are at least partially accounted for by other traumas and stressors, even though upon CAPS administration, study clinicians specifically inquired about WTC-related PTSD symptoms. Conversely, the high-risk nature of these individuals’ occupations may also mean increased exposure to resilience training for a sizable subsample, and greater access to social support networks of peers with similar experiences, potentially providing protective mechanisms.

In conclusion, this study has identified a vast number of biomarkers that will be potentially useful tools after independent validation. In particular, ten of these genes stand out as reproducible across multiple studies and should be considered as high priority. In combination with pathway and cellular deconvolution results, these findings highlight a strong connection with immune dysregulation and other psychiatric illnesses. We believe that future studies should focus on validation of our PTSD-associated genes and also single-cell RNA-sequencing approaches to delineate the role of immune cell types in PTSD.

## Data Availability

Gene expression summary statistics will be made available upon request.

---

Supplemental Table 2. (A) **Correlations of beta values and FDR-adjusted p values for CAPS lifetime versus past-month and all associated symptom dimensions. (B) Correlations of symptom dimensions against all other symptom dimensions for past-month and (C) lifetime.** All correlations were significant (p<2.2×10^-16^).

## References

1. Spitzer RL, Gibbon M, Williams JBW. Structured Clinical Interview for DSM-IV Axis I Disorders (SCID). N. Y. State Psychiatr. Inst. Biom. Res. N. Y. (1995).

2. Yehuda, R. et al. Gene Expression Patterns Associated with Posttraumatic Stress Disorder Following Exposure to the World Trade Center Attacks. Biol. Psychiatry 66, 708–711 (2009).

3. Kuan, P.-F. et al. Gene expression associated with PTSD in World Trade Center responders: An RNA sequencing study. Transl. Psychiatry 7, 1297 (2017).

4. Clouston, S. et al. Traumatic exposures, posttraumatic stress disorder, and cognitive functioning in World Trade Center responders. Alzheimers Dement. N. Y. N 3, 593–602 (2017).

5. Clouston, S. A. P. et al. Posttraumatic stress disorder and total amyloid burden and amyloid-β 42/40 ratios in plasma: Results from a pilot study of World Trade Center responders. Alzheimers Dement. Amst. Neth. 11, 216–220 (2019).

6. Clouston, S. A. P. et al. Incidence of mild cognitive impairment in World Trade Center responders: Long-term consequences of re-experiencing the events on 9/11/2001. Alzheimers Dement. Diagn. Assess. Dis. Monit. 11, 628–636 (2019).

7. Gong, Y. et al. Prostate Cancer in World Trade Center Responders Demonstrates Evidence of an Inflammatory Cascade. Mol. Cancer Res. 17, 1605–1612 (2019).

8. Kuan, P.-F. et al. Cell type-specific gene expression patterns associated with posttraumatic stress disorder in World Trade Center responders. Transl. Psychiatry 9, 1–11 (2019).

9. Kuan, P.-F. et al. Enhanced exposure assessment and genome-wide DNA methylation in World Trade Center disaster responders. Eur. J. Cancer Prev. 28, 225–233 (2019).

10. Sarapas, C. et al. Genetic Markers for PTSD Risk and Resilience Among Survivors of the World Trade Center Attacks. Dis. Markers 30, 101–110 (2011).

11. Huckins, L. M. et al. Analysis of Genetically Regulated Gene Expression identifies a trauma type specific PTSD gene, SNRNP35. bioRxiv 581124 (2019) doi:10.1101/581124.

12. Breen, M. S. et al. PTSD Blood Transcriptome Mega-Analysis: Shared Inflammatory Pathways across Biological Sex and Modes of Trauma. Neuropsychopharmacology 43, 469– 481 (2018).

13. Blake DD, Weathers FW, Nagy LM, et al. The development of a Clinician-Administered PTSD Scale. J Trauma Stress 8(1), 75–90 (1995).

14. Fanous, A. H. & Kendler, K. S. Genetic heterogeneity, modifier genes, and quantitative phenotypes in psychiatric illness: searching for a framework. Mol. Psychiatry 10, 6–13 (2005).

15. Schijven, D., et al. Multivariate genome-wide analysis of stress-related quantitative phenotypes. Eur. Neuropsychopharmacol. J. Eur. Coll. Neuropsychopharmacol. 29, 1354–1364 (2019).

16. Lee, S. H. & Wray, N. R. Novel Genetic Analysis for Case-Control Genome-Wide Association Studies: Quantification of Power and Genomic Prediction Accuracy. PLoS ONE 8, (2013).

17. Dasaro, C. R. et al. Cohort Profile: World Trade Center Health Program General Responder Cohort. Int. J. Epidemiol. 46, e9 (2017).

18. Ruggiero, K. J., Del Ben, K., Scotti, J. R. & Rabalais, A. E. Psychometric properties of the PTSD Checklist-Civilian Version. J. Trauma. Stress 16, 495–502 (2003).

19. Huckins, L. M. et al. Polygenic regulation of PTSD severity and outcomes among World Trade Center responders. medRxiv 2020.12.06.20244772 (2020) doi:10.1101/2020.12.06.20244772.

20. Bernstein, D. P. et al. Development and validation of a brief screening version of the Childhood Trauma Questionnaire. Child Abuse Negl. 27, 169–190 (2003).

21. Kubany, E. S. et al. Development and preliminary validation of a brief broad-spectrum measure of trauma exposure: the Traumatic Life Events Questionnaire. Psychol. Assess. 12, 210–224 (2000).

22. North, CS., et al. The Diagnostic Interview Schedule/Disaster Supplement (DIS-IV/DS). St Louis Wash. Univ. (2001).

23. Pietrzak, R. H., Goldstein, R. B., Southwick, S. M. & Grant, B. F. Medical comorbidity of full and partial posttraumatic stress disorder in US adults: results from Wave 2 of the National Epidemiologic Survey on Alcohol and Related Conditions. Psychosom. Med. 73, 697–707 (2011).

24. Horn, S. R. et al. Latent typologies of posttraumatic stress disorder in World Trade Center responders. J. Psychiatr. Res. 83, 151–159 (2016).

25. Pietrzak, R. H., Tsai, J., Harpaz-Rotem, I., Whealin, J. M. & Southwick, S. M. Support for a novel five-factor model of posttraumatic stress symptoms in three independent samples of Iraq/Afghanistan veterans: a confirmatory factor analytic study. J. Psychiatr. Res. 46, 317– 322 (2012).

26. Shah, H., Wang, Y.-C., Castellanos, R., Pandya, C., & Giles, Z. RAPiD - An Agile and Dependable RNA-Seq Framework. ASHG 2015 (2015).

27. Hoffman, G. E. & Schadt, E. E. variancePartition: interpreting drivers of variation in complex gene expression studies. BMC Bioinformatics 17, 483 (2016).

28. Law, C. W., Chen, Y., Shi, W. & Smyth, G. K. voom: precision weights unlock linear model analysis tools for RNA-seq read counts. Genome Biol. 15, R29 (2014).

29. Mellon, S. H., Gautam, A., Hammamieh, R., Jett, M. & Wolkowitz, O. M. Metabolism, Metabolomics, and Inflammation in Posttraumatic Stress Disorder. Biol. Psychiatry 83, 866– 875 (2018).

30. Wisnivesky, J. P. et al. Persistence of multiple illnesses in World Trade Center rescue and recovery workers: a cohort study. Lancet Lond. Engl. 378, 888–897 (2011).

31. Benjamini, Y. & Hochberg, Y. Controlling the False Discovery Rate: A Practical and Powerful Approach to Multiple Testing. J. R. Stat. Soc. Ser. B Methodol. 57, 289–300 (1995).

32. Bild, A. & Febbo, P. G. Application of a priori established gene sets to discover biologically important differential expression in microarray data. Proc. Natl. Acad. Sci. 102, 15278–15279 (2005).

33. Korotkevich, G., Sukhov, V. & Sergushichev, A. Fast gene set enrichment analysis. bioRxiv 060012 (2019) doi:10.1101/060012.

34. How to do GSEA in R with (fgsea or gage) and plot results. Bioinformatics Breakdown https://bioinformaticsbreakdown.com/how-to-gsea/ (2019).

35. Kanehisa, M., Sato, Y., Kawashima, M., Furumichi, M. & Tanabe, M. KEGG as a reference resource for gene and protein annotation. Nucleic Acids Res. 44, D457–D462 (2016).

36. Yu, G., Wang, L.-G., Han, Y. & He, Q.-Y. clusterProfiler: an R Package for Comparing Biological Themes Among Gene Clusters. OMICS J. Integr. Biol. 16, 284–287 (2012).

37. Willer, C. J., Li, Y. & Abecasis, G. R. METAL: fast and efficient meta-analysis of genomewide association scans. Bioinformatics 26, 2190–2191 (2010).

38. Newman, A. M. et al. Robust enumeration of cell subsets from tissue expression profiles. Nat. Methods 12, 453–457 (2015).

39. Daskalakis, N. P., Rijal, C. M., King, C., Huckins, L. M. & Ressler, K. J. Recent Genetics and Epigenetics Approaches to PTSD. Curr. Psychiatry Rep. 20, 30 (2018).

40. Neigh, G. N. & Ali, F. F. Co-Morbidity of PTSD and Immune System Dysfunction: Opportunities for Treatment. Curr. Opin. Pharmacol. 29, 104–110 (2016).

41. Wang, Z., Caughron, B. & Young, M. R. I. Posttraumatic Stress Disorder: An Immunological Disorder? Front. Psychiatry 8, (2017).

42. Speer, K. E., Semple, S., Naumovski, N., D’Cunha, N. M. & McKune, A. J. HPA axis function and diurnal cortisol in post-traumatic stress disorder: A systematic review. Neurobiol. Stress 11, (2019).

43. Yehuda, R., et al. Post-traumatic stress disorder. Nat. Rev. Dis. Primer 1, 1–22 (2015).

44. Nutt, D. J. & Malizia, A. L. Structural and functional brain changes in posttraumatic stress disorder. J. Clin. Psychiatry 65 Suppl 1, 11–17 (2004).

45. Duncan, L. E. et al. Largest GWAS of PTSD (N=20070) yields genetic overlap with schizophrenia and sex differences in heritability. Mol. Psychiatry 23, 666–673 (2018).

46. Li, Z. et al. Genome-wide association analysis identifies 30 new susceptibility loci for schizophrenia. Nat. Genet. 49, 1576–1583 (2017).

47. Jong, S. de et al. Immune signatures and disorder-specific patterns in a cross-disorder gene expression analysis. Br. J. Psychiatry 209, 202–208 (2016).

48. Tsolakidou, A. et al. Gene expression profiling in the stress control brain region hypothalamic paraventricular nucleus reveals a novel gene network including Amyloid beta Precursor Protein. BMC Genomics 11, 546 (2010).

49. Maccarrone, G. et al. Psychiatric patient stratification using biosignatures based on cerebrospinal fluid protein expression clusters. J. Psychiatr. Res. 47, 1572–1580 (2013).

50. Hou, Y., et al. Schizophrenia-associated rs4702 G allele-specific downregulation of FURIN expression by miR-338-3p reduces BDNF production. Schizophr. Res. 199, 176–180 (2018).

51. Schrode, N. et al. Synergistic effects of common schizophrenia risk variants. Nat. Genet. 51, 1475–1485 (2019).

52. English, J. A. et al. Reduced protein synthesis in schizophrenia patient-derived olfactory cells. Transl. Psychiatry 5, e663–e663 (2015).

53. Miron, J. et al. Association of PPP2R1A with Alzheimer’s disease and specific cognitive domains. Neurobiol. Aging 81, 234–243 (2019).

54. Bralten, J. et al. Candidate Genetic Pathways for Attention-Deficit/Hyperactivity Disorder (ADHD) Show Association to Hyperactive/Impulsive Symptoms in Children With ADHD. J. Am. Acad. Child Adolesc. Psychiatry 52, 1204–1212.e1 (2013).

55. Chang, W.-S., Wang, Y.-H., Zhu, X.-T. & Wu, C.-J. Genome-Wide Profiling of miRNA and mRNA Expression in Alzheimer’s Disease. Med. Sci. Monit. Int. Med. J. Exp. Clin. Res. 23, 2721–2731 (2017).

56. Kawai, T. et al. Gene expression signature in peripheral blood cells from medical students exposed to chronic psychological stress. Biol. Psychol. 76, 147–155 (2007).

57. Zhao, Y. et al. A large-scale integrative analysis of GWAS and common meQTLs across whole life course identifies genes, pathways and tissue/cell types for three major psychiatric disorders. Neurosci. Biobehav. Rev. 95, 347–352 (2018).

58. Konno, T. et al. DCTN1-related neurodegeneration: Perry syndrome and beyond. Parkinsonism Relat. Disord. 41, 14–24 (2017).

59. Seth, A., Giunta, S., Franceschil, C., Kola, I. & Venanzoni, M. C. Regulation of the human stress response gene GADD153 expression: role of ETS1 and FLI-1 gene products. Cell Death Differ. 6, 902–907 (1999).

60. St-Louis, É. et al. Involvement of the coatomer protein complex I in the intracellular traffic of the delta opioid receptor. Mol. Cell. Neurosci. 79, 53–63 (2017).

61. Bettayeb, K. et al. Relevance of the COPI complex for Alzheimer’s disease progression in vivo. Proc. Natl. Acad. Sci. U. S. A. 113, 5418–5423 (2016).

62. Yang, Y., Wang, X., Ju, W., Sun, L. & Zhang, H. Genetic and Expression Analysis of COPI Genes and Alzheimer’s Disease Susceptibility. Front. Genet. 10, 866 (2019).

63. Izumi, K. et al. ARCN1 Mutations Cause a Recognizable Craniofacial Syndrome Due to COPI-Mediated Transport Defects. Am. J. Hum. Genet. 99, 451–459 (2016).

64. Watkin, L. B. et al. COPA mutations impair ER-Golgi transport and cause hereditary autoimmune-mediated lung disease and arthritis. Nat. Genet. 47, 654–660 (2015).

65. Jean, F., Stuart, A. & Tarailo-Graovac, M. Dissecting the Genetic and Etiological Causes of Primary Microcephaly. Front. Neurol. 11, (2020).

66. Lippmann, M., Cohen, M. D. & Chen, L.-C. Health effects of World Trade Center (WTC) Dust: An unprecedented disaster with inadequate risk management. Crit. Rev. Toxicol. 45, 492– 530 (2015).

67. Reibman, J. et al. Characteristics of a Residential and Working Community With Diverse Exposure to World Trade Center Dust, Gas, and Fumes. J. Occup. Environ. Med. Am. Coll. Occup. Environ. Med. 51, 534–541 (2009).

68. Hervé, M. et al. Translational Identification of Transcriptional Signatures of Major Depression and Antidepressant Response. Front. Mol. Neurosci. 10, (2017).

69. Carlini, V. et al. CLIC1 Protein Accumulates in Circulating Monocyte Membrane during Neurodegeneration. Int. J. Mol. Sci. 21, (2020).

70. Tang, T. et al. CLICs-dependent chloride efflux is an essential and proximal upstream event for NLRP3 inflammasome activation. Nat. Commun. 8, 202 (2017).

71. Gurunathan, G., Yu, Z., Coulombe, Y., Masson, J.-Y. & Richard, S. Arginine methylation of hnRNPUL1 regulates interaction with NBS1 and recruitment to sites of DNA damage. Sci. Rep. 5, 10475 (2015).

72. Baker, K. et al. Neonatal Fc receptor for IgG (FcRn) regulates cross-presentation of IgG immune complexes by CD8−CD11b+ dendritic cells. Proc. Natl. Acad. Sci. U. S. A. 108, 9927– 9932 (2011).

73. Le-Niculescu, H. et al. Towards precision medicine for stress disorders: diagnostic biomarkers and targeted drugs. Mol. Psychiatry 25, 918–938 (2020).

74. Zhu, X., Bührer, C. & Wellmann, S. Cold-inducible proteins CIRP and RBM3, a unique couple with activities far beyond the cold. Cell. Mol. Life Sci. 73, 3839–3859 (2016).

75. Zhou, J.-Y. et al. Trauma-associated Human Neutrophil Alterations Revealed by Comparative Proteomics Profiling. Proteomics Clin. Appl. 7, (2013).

76. Lu, Q., Shen, N., Li, X. M. & Chen, S. L. Genomic view of IFN-α response in pre-autoimmune NZB/W and MRL/lpr mice. Genes Immun. 8, 590–603 (2007).

77. Chen, Y., Li, X., Kobayashi, I., Tsao, D. & Mellman, T. A. Expression and methylation in posttraumatic stress disorder and resilience; evidence of a role for odorant receptors. Psychiatry Res. 245, 36–44 (2016).

78. Breen, M. S. et al. Differential transcriptional response following glucocorticoid activation in cultured blood immune cells: a novel approach to PTSD biomarker development. Transl. Psychiatry 9, 201 (2019).

79. Glover, D. A., Steele, A. C., Stuber, M. L. & Fahey, J. L. Preliminary evidence for lymphocyte distribution differences at rest and after acute psychological stress in PTSD-symptomatic women. Brain. Behav. Immun. 19, 243–251 (2005).

80. Lemieux, A., Coe, C. L. & Carnes, M. Symptom severity predicts degree of T cell activation in adult women following childhood maltreatment. Brain. Behav. Immun. 22, 994–1003 (2008).

81. Skapenko, A., Leipe, J., Lipsky, P. E. & Schulze-Koops, H. The role of the T cell in autoimmune inflammation. Arthritis Res. Ther. 7, S4–S14 (2005).

82. Baumeister, D., Akhtar, R., Ciufolini, S., Pariante, C. M. & Mondelli, V. Childhood trauma and adulthood inflammation: a meta-analysis of peripheral C-reactive protein, interleukin-6 and tumour necrosis factor-α. Mol. Psychiatry 21, 642–649 (2016).

83. Dong, Y. et al. Stress-induced NLRP3 inflammasome activation negatively regulates fear memory in mice. J. Neuroinflammation 17, 205 (2020).

84. Fonkoue, I. T. et al. Symptom severity impacts sympathetic dysregulation and inflammation in post-traumatic stress disorder (PTSD). Brain. Behav. Immun. 83, 260–269 (2020).

85. Michopoulos, V., Powers, A., Gillespie, C. F., Ressler, K. J. & Jovanovic, T. Inflammation in Fear- and Anxiety-Based Disorders: PTSD, GAD, and Beyond. Neuropsychopharmacology 42, 254–270 (2017).

86. Speer, K., Upton, D., Semple, S. & McKune, A. Systemic low-grade inflammation in post-traumatic stress disorder: a systematic review. J. Inflamm. Res. 11, 111–121 (2018).

